# The impact of theta-burst stimulation on cortical GABA and glutamate in treatment-resistant depression: A surface-based MRSI analysis approach

**DOI:** 10.1101/2022.02.17.22271118

**Authors:** B Spurny-Dworak, GM Godbersen, MB Reed, J Unterholzner, T Vanicek, P Baldinger-Melich, A Hahn, GS Kranz, W Bogner, R Lanzenberger, S Kasper

**Affiliations:** Department of Psychiatry and Psychotherapy, Medical University of Vienna, Austria; Department of Rehabilitation Sciences, The Hong Kong Polytechnic University, Hung Hom, Hong Kong; Department of Biomedical Imaging and Image-guided Therapy, High Field MR Centre, Medical University of Vienna, Austria; Department of Molecular Neuroscience, Center for Brain Research, Medical University of Vienna, Austria

**Keywords:** TBS, MRS, GABA, glutamate, depression, TMS

## Abstract

**Background:** Theta burst stimulation (TBS) belongs to one of the biological antidepressant treatment options. When applied bilaterally, excitatory intermittent TBS (iTBS) is commonly targeted to the left and inhibitory continuous TBS (cTBS) to the right dorsolateral prefrontal cortex. TBS was shown to influence neurotransmitter systems, while iTBS is thought to interfere with glutamatergic circuits and cTBS to mediate GABAergic neurotransmission.

**Objectives:** We aimed to expand insights in the therapeutic effects of TBS on the GABAergic and glutamatergic system utilizing 3D-multivxovel magnetic resonance spectroscopy imaging (MRSI) in combination with a novel surface-based MRSI analysis approach to investigate changes of cortical neurotransmitter levels in patients with treatment resistant depression (TRD).

**Methods:** Twelve TRD patients (5 female, mean age±SD=35±11 years) completed paired MRSI measurements, using a GABA-edited 3D-multivoxel MEGA-LASER sequence, before and after three weeks of bilateral TBS treatment. Changes in cortical distributions of GABA+/tNAA (GABA+macromolecules relative to total N-acetylaspartate) and Glx/tNAA (Glx=mixed signal of glutamate and glutamine), were investigated in a surface-based region-of-interest (ROI) analysis approach.

**Results:** ANCOVAs revealed a significant increase in Glx/tNAA ratios in the left caudal middle frontal area (p_corr_.=0.046, F=13.292), an area targeted by iTBS treatment. Whereas, contralateral treatment with cTBS evoked no alterations in glutamate or GABA concentrations.

**Conclusion:** This study demonstrates surface-based adaptions in the stimulation area to the glutamate metabolism after excitatory iTBS but not after cTBS, using a novel surface-based analysis of 3D-MRSI data. The reported impact of facilitatory iTBS on glutamatergic neurotransmission provides further insight into the neurobiological effects of TBS in TRD.

## 1. Introduction

Major depressive disorder (MDD) represents a severe psychiatric disease affecting up to 3.8% of the population worldwide and has risen further during the last years (Collaborators, 2021). Several treatment options of pharmacological (e.g. selective serotonin reuptake inhibitors (SSRIs) or ketamine) or non-pharmacological, biological interventions (i.e. transcranial magnetic stimulation (TMS) or electroconvulsive therapy (ECT)) are currently available. Modifications of neurotransmitter systems are key aspects in the antidepressant actions of different interventions in order to restore GABAergic or glutamatergic function (Kalueff and Nutt, 2007; Sanacora, 2012). Several studies have shown SSRIs or ketamine to affect a variety of neurotransmitter systems including the serotonergic (Spindelegger, 2009; Hahn, 2010; Lanzenberger, 2012), GABAergic (Sanacora, 2002; Brennan, 2017; Silberbauer, 2020) or the glutamatergic system (Rowland, 2005; Taylor, 2008; Spurny, 2021). Moreover, certain antidepressants directly interfere with the glutamatergic or GABAergic system. The N-methyl-D-aspartate (NMDA) receptor antagonist ketamine is a treatment option for the use in TRD patients, leading to rapid symptom reductions (Kasper, 2021; McIntyre, 2021). Although ketamine is targeting the glutamatergic system, adaptions in GABA levels could be reported (Silberbauer, 2020). According to a recent study, the clinical efficacy in TRD of rTMS does not differ from ketamine (Mikellides, 2021). Another promising treatment option for TRD (currently under clinical investigation) is zuranolone a modulator of the GABA_A_ receptor (Gunduz-Bruce, 2019). Hence, both the glutamatergic and GABAergic system are promising targets for the treatment of TRD.

While the biological binding sites and downstream effects of pharmacological interventions are abundantly studied, this is oftentimes less clear for non-pharmacological, biological treatments. ECT is known to induce a remodeling of functional networks (Qi, 2020) and alterations across the serotonergic and GABAergic system (Sanacora, 2003; Baldinger, 2014).

Since TBS has fewer side effects compared to ECT, it finds broader acceptance in patients especially in treatment resistant depression (TRD). When two different pharmacological treatment trials fail to significantly improve clinical symptoms, MDD is commonly classified as TRD, although this definition varies between studies (Gaynes, 2020). In a meta-analysis of 29 randomized, double-blind and sham-controlled trials, Berlim et al. demonstrated response and remission rates of 29% and 19% of subjects with major depression receiving excitatory high-frequency TMS (Berlim, 2014). Thereby, typically bihemispheric theta burst stimulation (TBS) is applied using excitatory intermittent (iTBS) or inhibitory continuous TBS (cTBS) to the dorsolateral prefrontal cortex (DLPFC). The DLPFC was shown to provide a suitable target for TBS to treat TRD (George, 1995).

Previous imaging studies reported diverse effects of TMS on different morphological and physiological parameters. Stimulation of the DLPFC was reported to affect functional connectivity between the PFC and cingulate regions (Baeken, 2014; Salomons, 2014). Similar to pharmacological treatments, TMS was shown to evoke changes in neurotransmitter systems in both animal and human studies. Two ^1^H-MRS studies found correlations of glutamate levels in the motor cortex and excitability with TMS (Stagg, 2009; Tremblay, 2013). In disease, a study by Pogarell et al. revealed adaptions in the dopaminergic system in MDD patients following repetitive TMS (rTMS) treatment (Pogarell, 2006). Furthermore, an investigation by Lewis et al. reported changes in cortical excitability in patients suffering from MDD in the primary motor cortex and the ACC (Lewis, 2016). Dubin et al. were one of the first to investigate the therapeutic effect of TMS on neurotransmitter distribution using MRS, showing elevated GABA levels (Dubin, 2016). However, effects on the glutamatergic system in MDD are less conclusive and similar to GABA limited to a handful studies. While Dubin and colleagues reported no effects on glutamate in the PFC, a different approach revealed elevations in the glutamate/glutamine (Glu/Gln) ratio after TMS treatment in MDD patients (Croarkin, 2016; Dubin, 2016).

However, impacts on neurotransmitter levels seem to differ between intermittent and continuous TBS with studies reporting inconclusive findings. Iwabuchi et al. showed reduced GABA/Glx levels in the DLPFC and ACC after iTBS to the left DLPFC (Iwabuchi, 2017). Moreover, increased GABA concentrations in the PCC after iTBS to the left inferior parietal lobe could be revealed (Vidal-Pineiro, 2015). In addition iTBS was described to increase the N100 amplitude (a marker for GABA-mediated inhibition), while cTBS reduced this amplitude in the cerebellum of healthy individuals (Harrington and Hammond-Tooke, 2015). On the other hand, cTBS, applied to the motor cortex, was reported to increase GABA concentrations while no effects were shown on glutamate (Stagg, 2009) suggesting an enhancement of interneural circuits. Based on the evidence of preclinical and clinical studies in animals and humans, Li et al. proposed a model to explain differential aftermaths of intermittent and continuous TBS on GABA and glutamate (Li, 2019). This model suggests iTBS to inhibit GABAergic interneurons, which in consequence leads to reduced inhibition of glutamatergic pyramidal cells, while continuous bursts of cTBS might increase the inhibitory activities of interneurons resulting in higher GABA concentrations.

However, due to the inconclusive evidence of the potential of TBS to restore disrupted GABAergic and glutamatergic neurotransmission in TRD subjects further research is needed. Since previous findings of MRS studies were restricted to a limited number of locations by the use of single voxel sequences, we aimed to extend our understanding by applying a 3D-multivoxel MRSI approach to cover a range of cortical regions, involved in the pathophysiology of MDD. Due to the cortical stimulation method of TBS, changes within these regions are of high interest. Hence, we applied a novel surface-based analysis approach of multi-voxel MRS data, based on a similar method used in PET imaging (Greve, 2014). We investigated adaptions in cortical GABA+/tNAA (GABA+ = a combination of GABA and macromolecules; tNAA= total N-acetylaspartate) and Glx/tNAA (Glx = combined signal of glutamate and glutamine) after three weeks of iTBS to left and cTBS treatment to the right DLPFC in a cohort of TRD patients.

## 2. Methods

### 2.1. Study design

All study patients underwent three weeks of TBS treatment at the Department of Psychiatry and Psychotherapy at the Medical University of Vienna, Austria. MRSI measurements were conducted within 2 weeks prior to and after the TBS treatment period (see **Figure 1**). This study was approved by the Ethics Committee of the Medical University of Vienna (EK 1761/2015) and part of a larger clinical trial with multimodal neuroimaging (ClinicalTrials. gov Identifier: NCT02810717).

**Figure 1:**
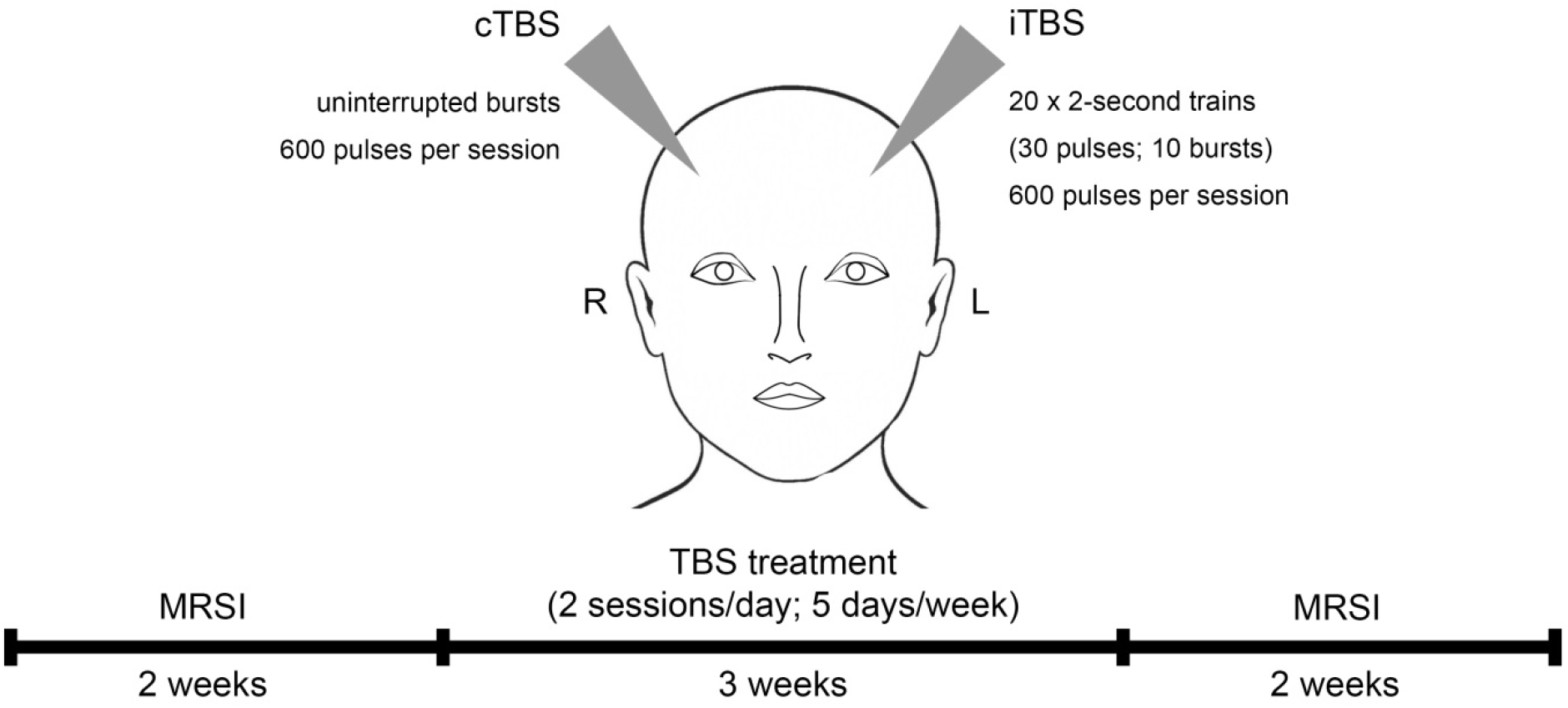
Study design and treatment regime. Study subjects received theta burst stimulation (TBS) over 3 weeks including 2 stimulations a day for 5 days per week. TBS sessions comprised intermittent TBS (iTBS) to the left and continuous TBS (cTBS) to the right dorsolateral prefrontal cortex. Magnetic resonance spectroscopy imaging (MRSI) measurements were conducted within 2 weeks before and after the treatment period.

### 2.2. Participants

Twelve TRD patients (5 female, mean age ± SD = 35 ± 11 years) with a DSM-4 diagnosis of single or recurrent MDD were included in our analysis. TRD was defined as insufficient response to two treatment trials in adequate dosage and time (> 4 weeks) according to the criteria set by the GSRD group (Group for the studies of Resistant Depression) (Bartova, 2019). Moreover, participants were included if they had a HAMD-17 total score of ≥ 18, a Clinical Global Impression Scale score of ≥ 4 and a stable treatment regime of 4 weeks prior to the study inclusion, which remained unchanged during the study participation. Exclusion criteria included psychotic symptoms, severe internal illnesses within the last 5 years, neurological diseases or brain injuries, substance abuse left handedness, or any contraindications to TMS treatment and MRI.

### 2.3. Transcranial magnetic stimulation

Over the course of three weeks, patients received intermittent (stimulating) TBS (iTBS) to the left DLPFC and continuous (inhibiting) TBS (cTBS) to the right DLPFC. iTBS consisted of 2-second trains (30 pulses; 10 bursts) repeated 20 times (600 pulses per session). cTBS comprised uninterrupted bursts of 600 pulses per session (see **Figure 1**). The TBS protocol was performed similar to Huang et al. (3-pulse 50-Hz bursts delivered at 5Hz) by using a MagPro magnetic stimulator (MagVenture, Denmark K) and a figure-of-eight shaped cool coil (Cool-B70) (Huang, 2005). Daily treatment (5 days per week) included two TBS sessions, separated by one hour, which were reversed for consecutive sessions (Li, 2014). The stimulation area (DLPFC) was defined in Montreal Neurological Institute (MNI) space (coordinates: [-38, +44, +26] - left DLPFC; [+38, +44, +26] - right DLPFC), using neuro-navigation (LOCALITE® TMS Navigator Germany), based on individual structural MRIs of each participant (Hecht, 2010). Stimulation intensity was based on 120% of the individual resting motor threshold (Ge, 2017).

### 2.4. Magnetic resonance spectroscopy

MRI measurements were performed on a 3 Tesla MAGNETOM Prisma Siemens MR Scanner using a 64-channel head coil. For accurate volume of interest (VOI)-placement and surface extraction, 3D T1-weighted anatomical images were acquired via a MPRAGE sequence (208 slices, 288×288 matrix size, voxel size 1.15×1.15×0.85mm^3^) with GRAPPA acceleration. For MRS, a constant-density, spiral-encoded, 3D-MRSI sequence with MEGA-LASER editing (Bogner, 2014) was used with a VOI = 110×120×45mm^3^ and field of view (FOV) = 160×160×160mm^3^. The acquired matrix size of 10×10×10 (approx. 4cm^3^ voxel size) was interpolated to a 16×16×16 matrix (approx. 1cm^3^ voxel size) during spectral processing steps. Since the VOI was placed close to the skull to cover cortical regions, tissue saturation slabs (25mm thickness, sat. delta frequency: -3.5 ppm) were used to suppress signals from subcutaneo jjus lipids (see **Figure 2**). Siemens advanced shimming procedure with manual adjustments was used. During the EDIT-ON acquisition, MEGA -editing pulses (60 Hz Gaussian pulses of 14.8 ms duration) were set to 1.9 ppm, editing the coupled 4CH_2_ triplet of GABA resonating at 3.02 ppm. 24 acquisition-weighted averages and two -step phase cycling were employed for 3D-MRSI, resulting in a total scan time of 17:23 min.

**Figure 2:**
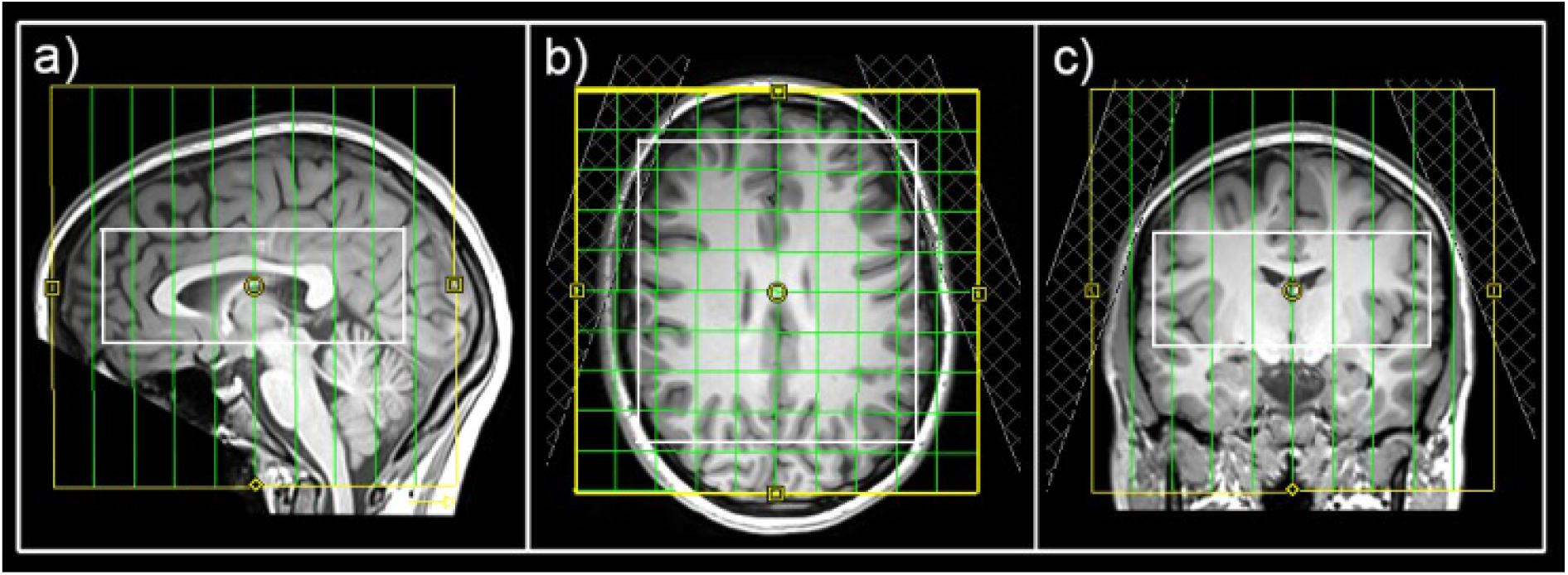
Placement of the field of view (yellow), volume of interest (white) and tissue separation slaps in sagittal (a), horizontal (b) and coronal (c) view.

An in-house software tool using MATLAB (R2013a, MathWorks, Natick, MA, USA), Bash (4.2.25, Free Software Foundation, Boston, MA, USA), MINC (2.0, MINC Tools, McConnell Brain Imaging Center, Montreal, QC, Canada) and LCModel software (6.3–1, S. Provencher, LCModel, Oakville, ON, Canada) was used for the quantification of all spectra within the VOI (Spurny, 2019). A simulated basis sets was created using the GAMMA library for the difference spectrum (containing GABA+, Glx and tNAA among others, (Hnilicova, 2016)). An exemplary spectrum is shown in **supplement figure 1**. Cramér– Rao lower bounds (CRLB) thresholds were set at 30% and spectra were visually inspected.

### 2.5. Surface-based MRSI analysis

For surface-based quantification, metabolic maps of GABA+, Glx and tNAA were interpolated to the resolution of anatomical images and ratio maps of GABA+/tNAA and Glx/tNAA were calculated. Ratios to tNAA were favored over total creatine (tCr), since changes in tCr after rTMS treatment were previously reported (Grohn, 2019). FreeSurfer 6.0 (https://surfer.nmr.mgh.harvard.edu/) was used for the surface-based analysis approach of MRSI data. Previous investigations have successfully shown cortical analysis approaches of metabolic maps in FreeSurfer using positron emission tomography (PET) data (Greve, 2014). Hence, this analysis was based on previous reports. Individual ratio maps of single subjects were spatially normalized by projecting onto the standard surface (fsaverage) using the tkregister2 command. All vertices of individual surfaces were assigned to the corresponding region-of-interest (ROI) using the Desikan atlas (Desikan, 2006). The following ROIs were included in the analysis: superiorfrontal, rostral middle frontal, caudal middle frontal, pars opercularis and precentral for both Glx/tNAA and GABA+/tCr and additionally pars triangularis, postcentral, paracentral, posterior cingulate and caudal anterior cingulate for Glx/tNAA ratios only, due to insufficient data quality in GABA+ maps. Furthermore, each surface was filtered by removing vertices that did not pass the CRLB threshold or laid above twice the standard variation within its respective brain region. After filtering steps, all remaining vertices were averaged within each ROI. Group-wise comparisons between measurements were done with calculated mean cortical neurotransmitter ratios within ROIs of each subject.

### 2.6. Statistical analysis

Statistical analyses were performed using SPSS Statistics (v26.0, 2010, SPSS, Inc., an IBM Company, Chicago, United States of America). Two-tailed paired t-tests were conducted to test for differences in HAM-D measures before and after the treatment period (p < 0.025). Univariate analyses of covariance (ANCOVAs) including sex and age as covariates were performed for each ROI and neurotransmitter ratio independently, to test for differences between measurements. Sidak correction was applied to correct for multiple comparisons (ROIs * neurotransmitter ratios, **Table 1**). Residuals were tested for normal distribution using the Kolmogorov-Smirnoff test.

**Table 1:**
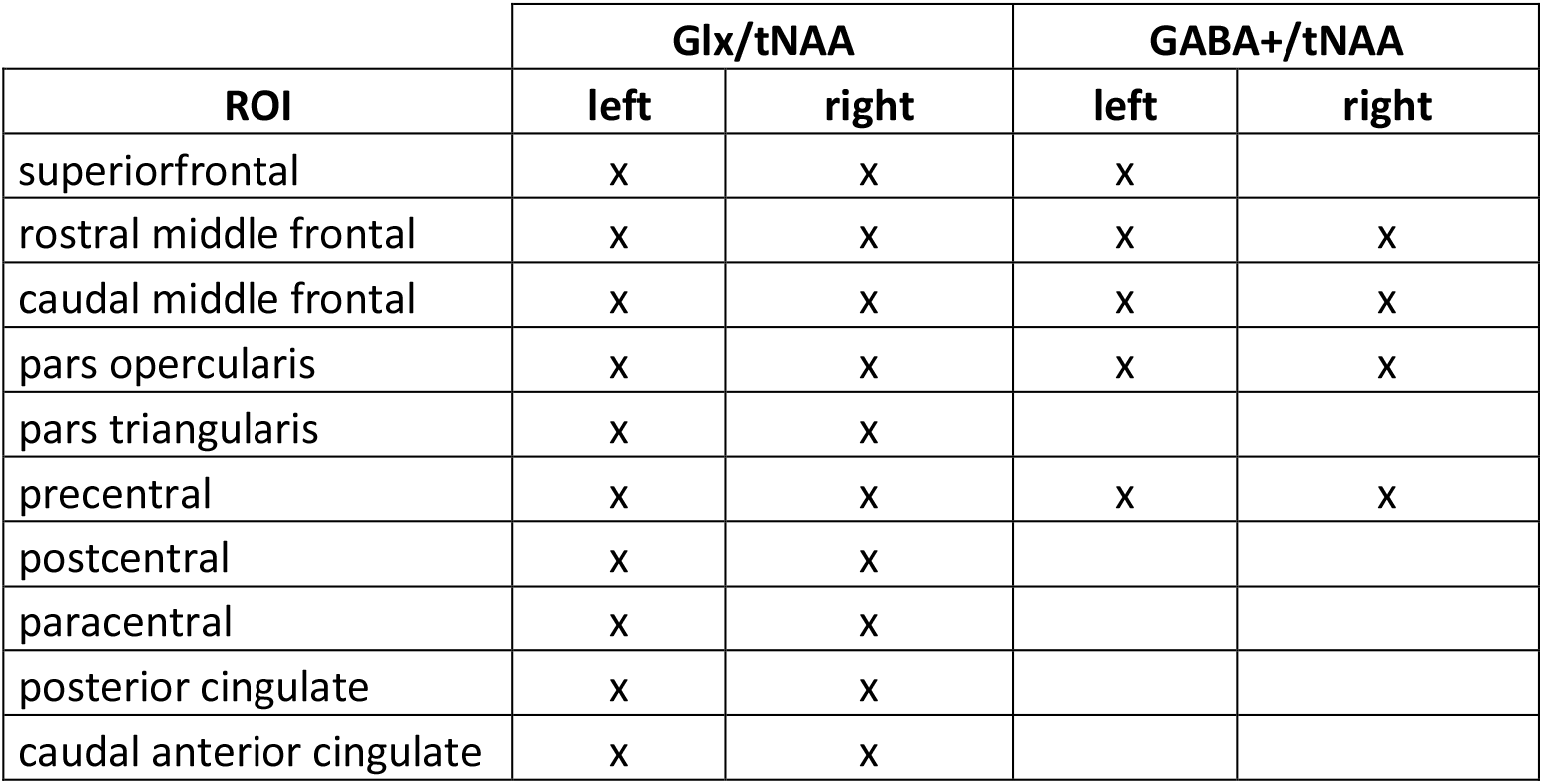
ROIs of each hemisphere included in the analysis of Glx/tNAA and GABA+/tNAA ratios. Glx = combined measure of glutamate and glutamine; tNAA = total N-acetylaspartate; GABA+ = a combination of GABA and macromolecules.

| ROI | Glx/tNAA |  | GABA+/tNAA |  |
| --- | --- | --- | --- | --- |
|  | left | right | left | right |
| superiorfrontal | x | x | x |  |
| rostral middle frontal | x | x | x | x |
| caudal middle frontal | x | x | x | x |
| pars opercularis | x | x | x | x |
| pars triangularis | x | x |  |  |
| precentral | x | x | x | x |
| postcentral | x | x |  |  |
| paracentral | x | x |  |  |
| posterior cingulate | x | x |  |  |
| caudal anterior cingulate | x | x |  |  |

## 3. Results

All twelve TRD patients (5 female, mean age ± SD = 35 ± 11 years) completed both MRSI measurements. Detailed stable pharmacological treatment of the patient cohort can be found in **supplement table 1**. HAM-D measures showed significant reductions after the treatment period (19.9 ± 2.8 before treatment to 12 ± 6.8 post treatment (mean ± SD), p.= 0.002) with a response rate of 33% (HAM-D reductions ≥ 50%) and remission rate of 25% (HAM-D < 7) of the TRD patients.

Due to insufficient data quality of GABA+ maps in the right superiofrontal area, this ROI had to be excluded resulting in a total of 29 ROIs in the final analysis (see **Table 1**). Hence, results from ANCOVAs were corrected for 29 comparisons using the Sidak correction method.

ANCOVAs revealed a significant difference in Glx/tNAA ratios in the left caudal middle frontal area (p_corr_. = 0.046, F = 13.292), an area targeted by (excitatory) iTBS treatment. Boxplots illustrating mean Glx/tNAA ratios before and after the treatment are shown in **figure 3**. No changes in GABA+/tNAA ratios could be detected in any ROI investigated. Although a cluster of elevated GABA+/tNAA ratios can be seen in the right caudal middle frontal area (see **Figure 3d**), an area targeted with inhibitory cTBS, changes within this area did not reach statistical significance.

**Figure 3:**
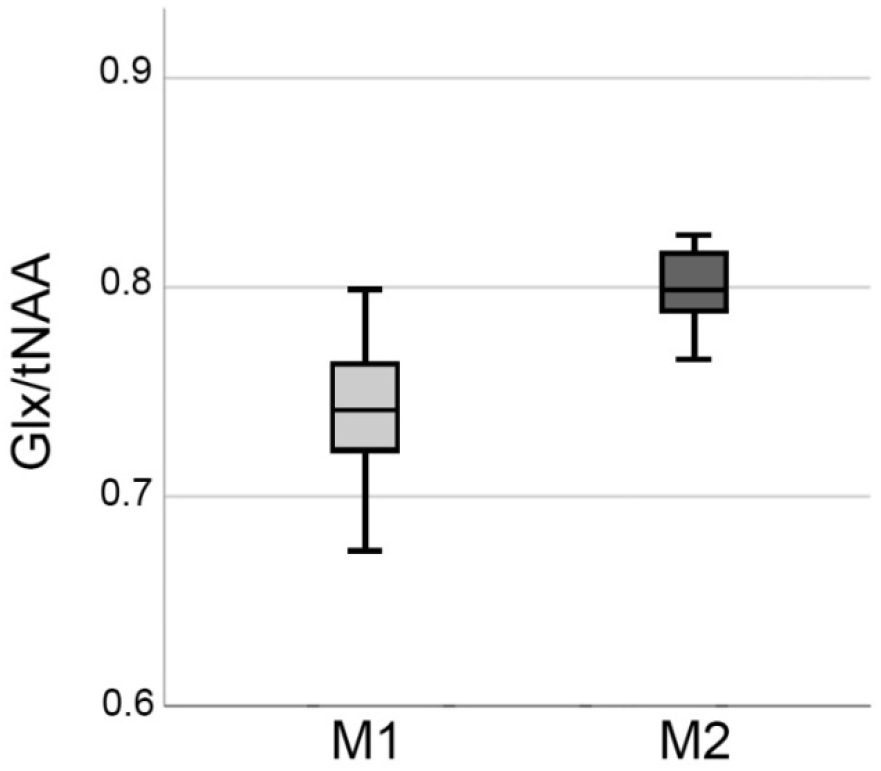
Boxplots showing elevations in Glx/tNAA ratio in the right caudal middle frontal area before (M1) and after the treatment period (M2). Glx = combined measure of glutamate and glutamine; tNAA = total N-acetylaspartate.

Changes in cortical Glx/tNAA and GABA+/tNAA are depicted in **figure 4**. Moreover, distributions of Glx/tNAA and GABA+/tNAA before and after the treatment are shown in **figure 5** and **supplement figure 2**.

**Figure 4:**
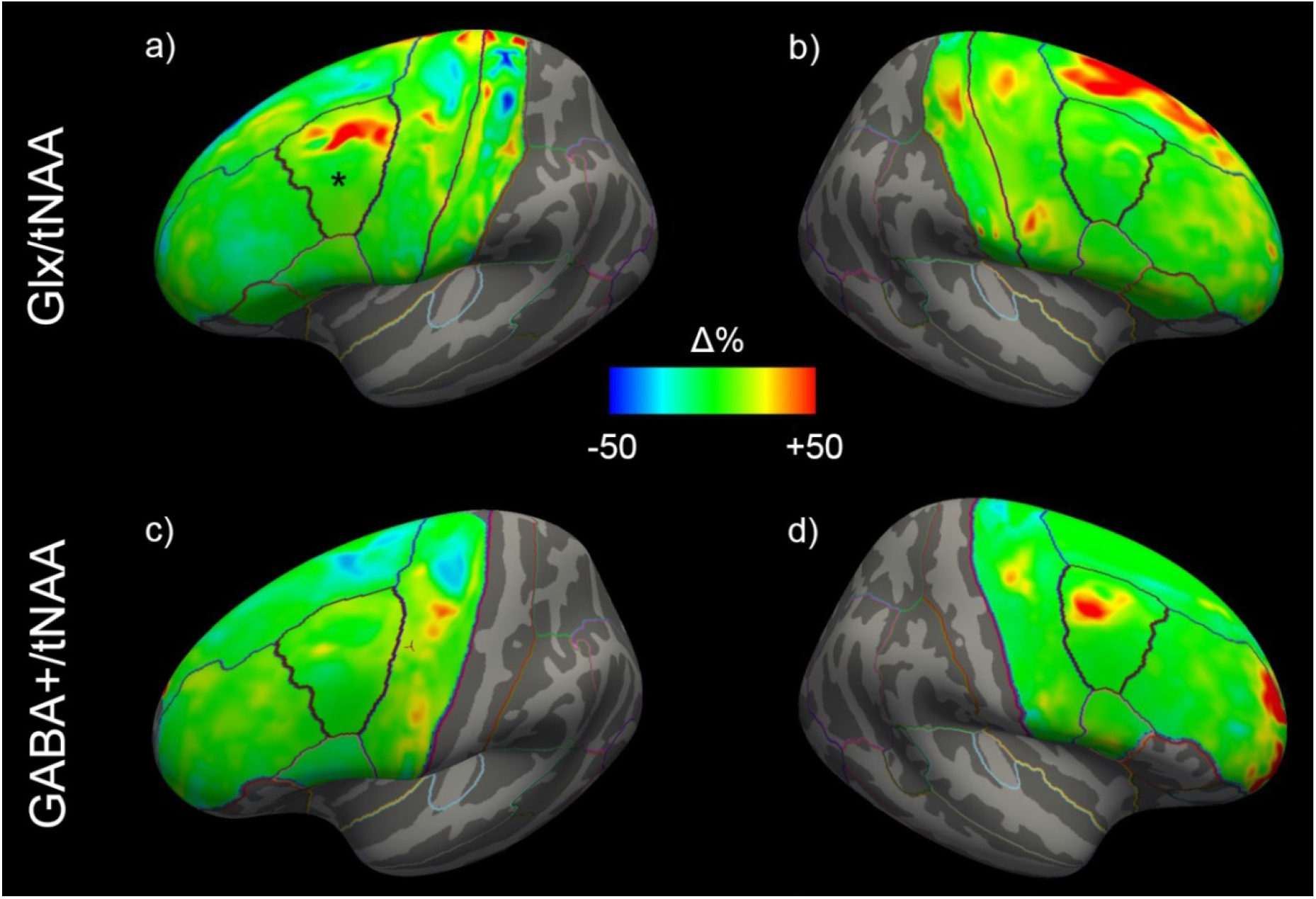
Mean changes of Glx/tNAA and GABA+/tNAA ratios of the left (a,c) and right (b,d) hemisphere across all study participants. The left caudal middle frontal area in (a), showing significant changes, is marked with a *. Glx = combined measure of glutamate and glutamine; tNAA = total N-acetylaspartate; GABA+ = a combination of GABA and macromolecules.

**Figure 5:**
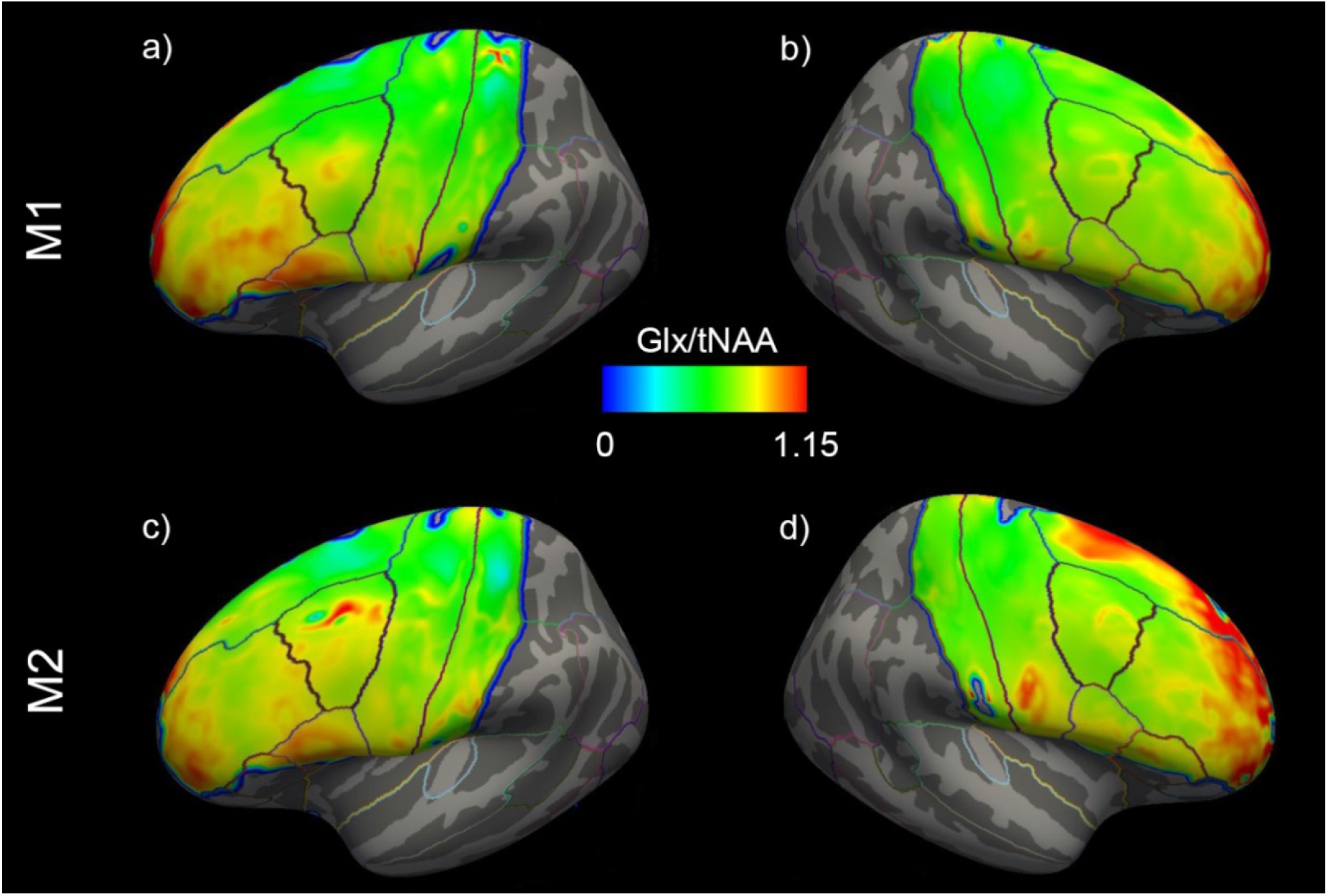
Mean distribution of Glx/tNAA ratios before (=M1) and after (=M2) the treatment period of the left (a and c) and right (b and d) hemisphere. Glx = combined measure of glutamate and glutamine; tNAA = total N-acetylaspartate.

## 4. Discussion

Here we report elevated Glx/tNAA ratios in the left caudal middle frontal area after 3 weeks of TBS treatment in TRD patients using a surface-based MRSI analysis approach. Significant increases of Glx/tNAA were found in the left caudal middle frontal area after iTBS, while Glx/tNAA in the corresponding right area remained unchanged after cTBS. No changes in GABA+/tNAA ratios were revealed across the investigated cortical regions. Similar to previous surface-based PET analysis approaches (Greve, 2014), the proposed surface-based investigations of MRSI data provides a suitable tool when adaptions in neurotransmitter levels of cortical regions are expected, i.e. by utilizing cortical stimulation methods. Moreover, following the treatment, TRD patients experienced a marked reduction of HAM-D scores, a response rate of ∼33%, and a remission rate of ∼25%, which seems promising when compared to the 13.7% remission rate of the equivalent TRD patient collective in the third treatment step of the STAR*D study (Rush, 2006). Hence, the impact of the stimulation on neurotransmitters gives further insight into the neurobiological effects of TBS.

A dysregulation of glutamate, glutamine and GABA metabolism in MDD could be previously shown (Croarkin, 2011; Sanacora, 2012; Abdallah, 2014). Moreover, rTMS was assumed to directly influence these neurotransmitter systems. In line with (Croarkin, 2016), showing elevated glutamine/glutamate ratios in the anterior cingulate cortex and DLPFC, we found an increase of Glx/tNAA levels in the stimulation area of iTBS. Prior studies suggested a modulation of the glutamatergic system after rTMS demonstrating higher glutamate levels in both preclinical (Yue, 2009) and clinical studies (Michael, 2003). Moreover, Luborzewski et al. were able to show a link between clinical effectiveness of rTMS treatment to the DLPFC and glutamate elevations through rTMS therapy (Luborzewski, 2007). On a neurobiological level, rTMS is thought to alter synaptic connections and thereby affecting long-term potentiation (Fitzgerald, 2006). Hence, cTBS is suggested to lower synaptic strength, while iTBS leads to opposite effects in both GABAergic and glutamatergic cells (Huang, 2005; Huang, 2007). These findings highlight the importance of glutamate and its receptors to the physiological TBS response in the human brain (Huang, 2007; Ishikawa, 2007; Li, 2019). As proposed in the model by Li and colleagues, describing the neurophysiological effects of TBS treatment, the stimulating or facilitatory iTBS is thought to be accompanied by inhibition of GABAergic interneurons via feedforward inhibition leading to decreased suppression of glutamatergic cells (Li, 2019). In consideration of this model, our results demonstrate increased Glx/tNAA ratios in the stimulation area of iTBS. Hence, the decreased suppression of glutamatergic cells seems to be reflected in increased Glx/tNAA content, while the inhibition of interneurons was not reflected in altered GABA+/tNAA ratios.

On the other hand, inhibitory cTBS is speculated to activiate the I-1 pathway as well as leading to long-term depression by prolonged Ca2+ increases and thereby slowly increasing the activity of GABAergic interneurons (Li, 2019). This was supported by increased GABA levels detected after cTBS to the motor cortex, without changes in glutamate being observed (Stagg, 2009). Moreover, several studies implicated a contribution of GABAergic neurotransmission in TBS-evoked plasticity (Larson and Munkacsy, 2015). Findings that cTBS lead to decreased numbers in calbindin interneurons (Suppa, 2016) or a report of a modulation of different classes of interneurons after both cTBS and iTBS (Labedi, 2014) adds to the importance of the interneural network in TBS mechanisms. Interestingly, increases in GABA concentrations could be shown after both iTBS and cTBS when some clinical studies reported elevated GABA levels or an influence on marker for GABAergic inhibition (Harrington and Hammond-Tooke, 2015; Vidal-Pineiro, 2015; Dubin, 2016) after iTBS treatment in patients, while also cTBS was shown to increase GABA concentrations in the motor cortex of healthy individuals (Stagg, 2009). However, the attribution of GABAergic interneurons could not be reflected in alterations in total GABA+ content in the scope of our study. Although, there seems to be a cluster with GABA+/tNAA increases in the stimulation area of cTBS, our data did not reach statistical significance within this ROI. Hence, in contrast to previous MRS studies of patient cohorts we could not show GABA alterations after iTBS (Harrington and Hammond-Tooke, 2015; Dubin, 2016).

This surface-based analysis approach for MRSI data provides a suitable method when changes in cortical neurotransmitter concentrations are of interest. Based on previous surface-based PET analyses approaches (Greve, 2014), analysis of cortical metabolites can be done using cortex-sbed atlases in individual subjects (Desikan, 2006). However, an appropriate MRSI sequence with reliable signal suppression in lipid-rich areas is required. Moreover, this method allows the quantification of several cortical regions simultaneously, which discriminates these investigations from previous MRS studies focusing on the effects of rTMS treatment using single voxel approaches in very selected brain regions.

Some limitations of this study need to be mentioned. While the use of GABA-edited MRSI provides the basis for the quantification of both excitatory and inhibitory neurotransmitters, the GABA signal is prone to artifacts. Hence, the data available was limited to a restricted number of cortical brain regions compared to derived Glx maps. All TRD patients included in these analyses had stable treatment regimens of at least 4 weeks prior to the study inclusion, which remained unchanged in the course of this study. However, an attribution of pharmacological interventions in the derived GABA+ and Glx concentrations cannot be excluded. Moreover, we could not include a group receiving sham treatment due to the limited number in available TRD patients undergoing MRSI.

## Conclusion

This study demonstrates a significant increase of Glx/tNAA ratios in the stimulation area of excitatory iTBS treatment of TRD patients. Our findings suggests changes in glutamate metabolism, following excitatory iTBS, to be mediated by reduced inhibition of pyramidal cell, while neurotransmitter concentrations remained stable after inhibitory cTBS on the contralateral hemisphere. These results may help to contribute to a better understanding of neurobiological implications of TBS in TRD patients.

## Data Availability

All data produced in the present study are available upon reasonable request to the authors

## 6. Acknowledgements

We thank Richard Frey, Gregor Gryglewski, Marius Hienert, Marie Spies, Christoph Kraus, Alexander Kautzky, Arkadiusz Komorowski, Paul Michenthaler and the diploma students of the Neuroimaging Labs (NIL) for medical support. Moreover, we would like to express our gratitude towards Sebastian Ganger, Philipp Moser and Eva Heckova for technical support, and all additional staff involved in the realization of this research.

## 8. Data availability statement

Due to data protection laws processed data is available from the authors upon reasonable request. Please contact with any questions or requests.

