## Supplement Material for "The impact of theta-burst stimulation on cortical GABA and glutamate in treatment-resistant depression: A surface-based MRSI analysis approach"

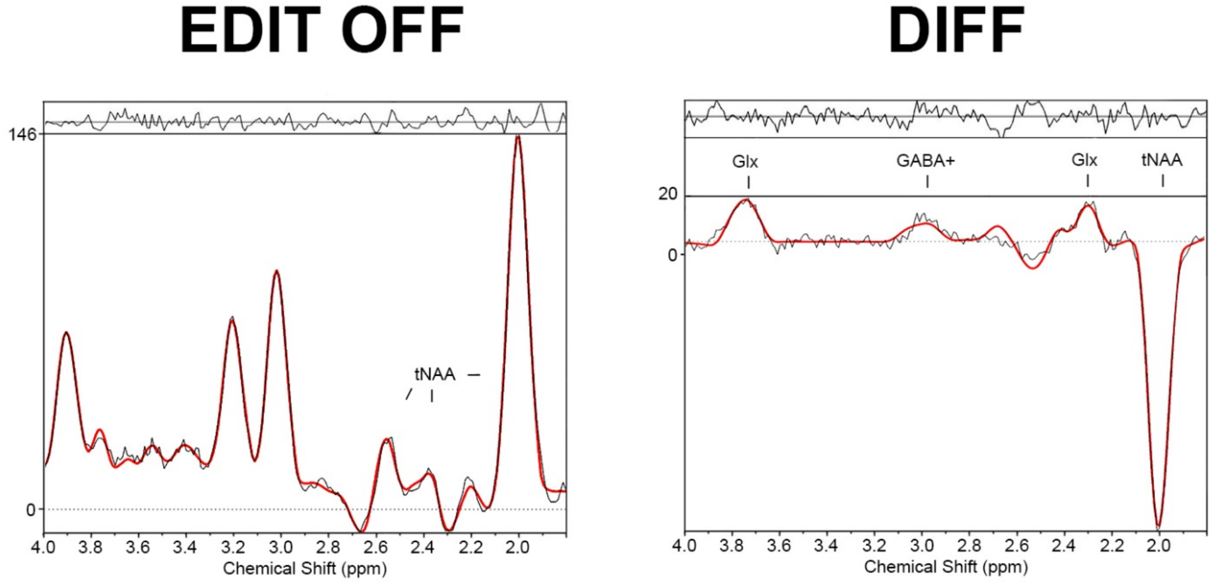


**Supplement figure 1:** Exemplary unedited (EDIT OFF) and edited (DIFF) spectra derived from the caudal middle frontal area showing peaks of total N-acetylaspartate (tNAA), a combination of GABA and macromolecules (=GABA+) and a combined signal of glutamate and glutamine (=Glx).

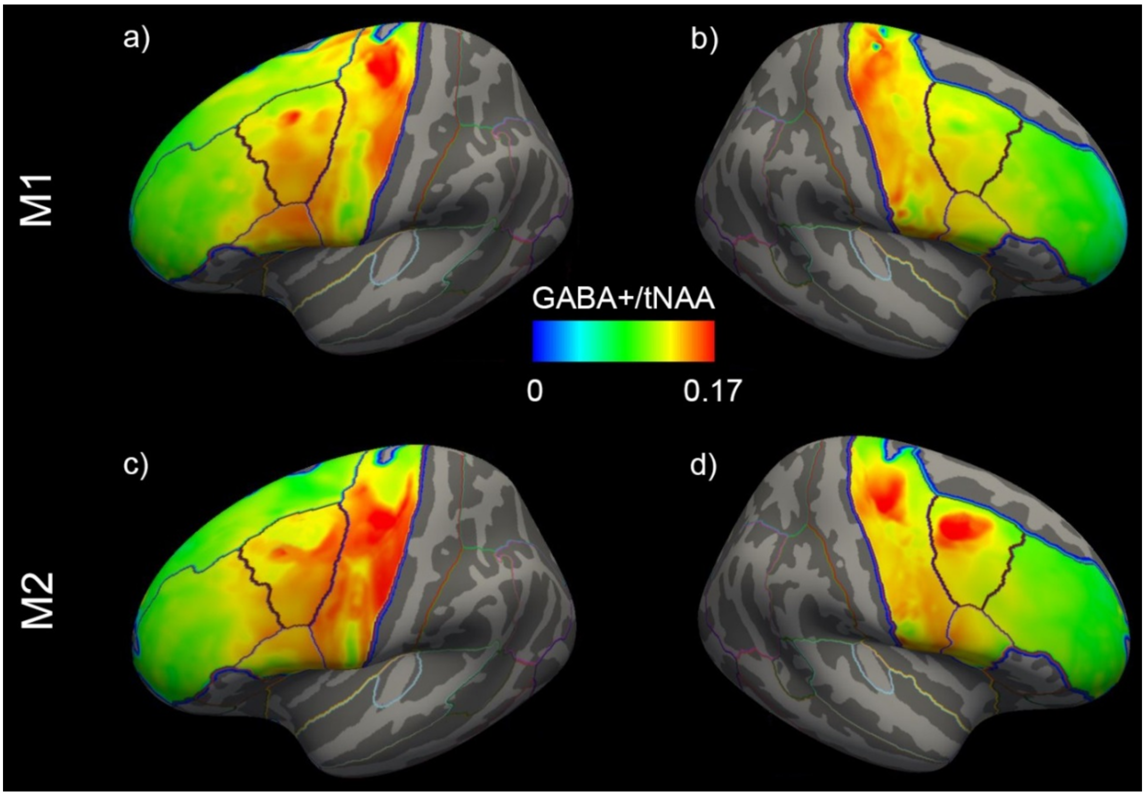


**Supplement figure 2:** Mean distribution of GABA+/tNAA ratios before (=M1) and after (=M2) the treatment period of the left (a and c) and right (b and d) hemisphere. tNAA = total N-acetylaspartate; GABA+ = a combination of GABA and macromolecules.

**Supplement table 1:** Detailed medication of the TRD patients (with dosages in milligram where documented)

| subject | medication |
|---|---|
| 1 | sertraline 100mg, pregabalin |
| 2 | venlafaxine 150mg, bupropion 150mg, lamotrigine 100mg, pregabalin 225mg, olanzapine 2.5mg, amitriptyline 20mg |
| 3 | none |
| 4 | sertraline 200mg, escitalopram 5mg |
| 5 | none |
| 6 | sertraline 100mg |
| 7 | escitalopram 10mg, mianserin 15mg |
| 8 | escitalopram 30mg |
| 9 | venlafaxine 150mg, lithium 450mg |
| 10 | milnacipran |
| 11 | melitracen |
| 12 | none |
